# COVID-CT-Mask-Net: Prediction of COVID-19 from CT Scans Using Regional Features

**DOI:** 10.1101/2020.10.11.20211052

**Authors:** Aram Ter-Sarkisov

## Abstract

We present COVID-CT-Mask-Net model that predicts COVID-19 from CT scans. The model works in two stages: first, it detects the instances of ground glass opacity and consolidation in CT scans, then predicts the condition from the ranked bounding box detections. To develop the solution for the three-class problem (COVID, common pneumonia and control), we used the COVIDx-CT dataset derived from the dataset of CT scans collected by China National Center for Bioinformation. We use about 5% of the training split of COVIDx-CT to train the model, and without any complicated data normalization, balancing and regularization, and training only a small fraction of the model’s parameters, we achieve a **90**.**80**% COVID sensitivity, **91**.**62**% common pneumonia sensitivity and **92**.**10**% normal sensitivity, and an overall accuracy of **91**.**66%** on the test data (21182 images), bringing the ratio of test/train data to **7**.**06**, which implies a very high capacity of the model to generalize to new data. We also establish an important result, that ranked regional predictions (bounding boxes with scores) in Mask R-CNN can be used to make accurate predictions of the image class. The full source code, models and pretrained weights are available on https://github.com/AlexTS1980/COVID-CT-Mask-Net.

## 1 Introduction

Deep learning COVID diagnostic tools for a 3 classes problem (COVID vs common pneumonia vs control) from CT images include COVIDNet-CT [GWW20], that consists of a single feature extractor trained on COVIDx-CT dataset split, COVNet (augmented ResNet50) [LQX^+^20] and ResNet18 [BGCB20]. Distinguishing between 3 classes is a more challenging problem than COVID vs Pneumonia or COVID vs Control, due to the larger number of potential false predictions. In order to achieve the state-of-the-art [GWW20] accuracy, large amounts of data are required to train (about 60K images) the model, that are often not available, which explains demand for various augmentations.

One approach that some publications use for COVID prediction, is the semantic segmentation model, e.g. U-Net in [ZLS^+^20, WGM^+^20] as a pre-processing step: its output (mask) is used by the classifier to enhance the prediction. The advantage of using a segmentation model is that it is capable of explicitly learning and predicting areas of infection associated with COVID. For a binary classification problem (COVID vs control, COVID vs common pneumonia), COVID-CT [ZZHX20] and JCS (Joint Classification and Segmentation) [WGM^+^20] are publicly available. COVID-CT combines lung masks predicted using U-Net with deep image features extracted using DenseNet169 and ResNet50 to predict the class, achieving an overall accuracy of 89% on the test data of about 350 images. JCS uses a similar approach, but with additional loss functions at deep layers (multiscale training), achieving a Dice score of 0.783 on the test data of about 120K images.

A number of reviews have compared different feature extractors and models directly to establish the best one for accuracy: [SZL^+^, SWS^+^20, LYZ^+^20]. From these papers it seems that for CT scans data, ResNet50 (+Feature Pyramid Network), ResNeXt and DenseNet121 produce the highest overall accuracy. At least one recent publication [ARK20] discusses the use of Mask R-CNN for predicting COVID from the segmentation of CT scans.

The majority of COVID deep learning models use radiography (X-rays) data due to its prevalence, e.g. the state-of-the-art COVID-Net [WW20] that has an architecture similar to COVIDNet-CT. To the best of our knowledge, only COVIDNet-CT, COVNet and ResNet18 [BGCB20] use CT scans for a 3-class (COVID vs common pneumonia vs control) rather than a binary (COVID vs control) problem. This problem is more challenging and realistic, both due to the fact that COVID and common pneumonia symptoms are similar in many ways, on CT scans they manifest in a different way [ZZX^+^20a, ZZX^+^20b, YWR^+^20], but these differences are subtle. These models have the following drawbacks: COVIDNet-CT requires a large training data with various augmentations and class balancing to achieve the state-of-the-art accuracy and COVID sensitivity, COVNet was evaluated on a small dataset (about 500 images), ResNet18 [BGCB20] is not publicly available, it has a low COVID sensitivity (81.2%) and was evaluated on a small data (90 images).

In this paper we would like to address these shortcomings by extending the semantic segmentation+classification solution to instance segmentation+classification using Mask R-CNN. Mask R-CNN [HGDG17] and Faster R-CNN [RHGS15] are the state-of-the-art models in instance segmentation and object detection. Mask R-CNN is an extension of Faster R-CNN with a mask prediction branch at an instance level. This is different to semantic segmentation models like Fully Convolutional Network (FCN) [LSD15] and U-Net [RFB15], which predict objects at pixel level. Mask R-CNN differentiates between different instances belonging to the same class by predicting their location (bounding box co-ordinates) using Region Proposal Network (RPN) and Regions of Interest (RoI). Each predicted object has therefore three features: bounding box, class and mask. The strength of Faster/Mask R-CNN comes from the fact that the model constructs samples of data from each image (regional features) to make predictions about the instances. This leverages the scarcity of the training data, and we use this strength both to obtain accurate predictions and use a small sample for training. By augmenting Mask R-CNN with a classification module, we extend its ability to detect objects to making prediction about the whole image.

The novelty of our approach to COVID-19 prediction can be summarized in the following way:

1. Solution: we use approx. 5% of the COVIDx-CT training data, and 3% of the total data, and, without any data augmentation, e.g. class weights, background removal and batch balancing, on which COVIDNet-CT depends, achieve 90.80% COVID sensitivity, and 91.66% overall accuracy on the full test split (21182 images).
2. Architecture: we repurpose Mask R-CNN to predict the class of the image using bounding box predictions by leveraging the ability of Mask R-CNN to extract regions of interest (RoIs) from deep features and obtain spatial predictions (bounding boxes) from them to construct a batch of ranked regional predictions in each image and use it to predict the global (image) class.
3. We solve both segmentation and prediction problems by training two models. Mask R-CNN segmentation model predicts and segments the cases of Ground Glass Opacity and Consolidation in CT scans, COVID-CT-Mask-Net uses this model to predict the class of the image.

Overall, we use much less training data than COVIDNet-CT, achieve a better overall accuracy and COVID sensitivity than COVNet[ZZX^+^20a] and ResNet18[BGCB20], and our solution has a better potential for generalization to other datasets.

## 2 Data

### 2.1 Segmentation

For our segmentation model we use the publicly available dataset published by China National Center for Bioinformation [ZLS^+^20] consisting of 650 scans across 150 patients with various stages of COVID. A total of 1+2 classes are segmented at pixel level: lung field (normal), which we merged with the background, ground glass opacity (GGO) and consolidation (C). These two conditions are often associated with various stages of COVID and other viral diseases, so we treat them as positive classes. We randomly split the provided dataset into 500 training and 150 validation images, maintaining the patient consistency, therefore some slices of COVID-positive patients do not contain positive classes. The challenge of the data is summarized in Figure 1: it is clear that positive scans can contain a small number of small objects of either class, and overall, the proportion of positive areas is very low, making the problem of segmenting them a serious challenge.

**Figure 1:**
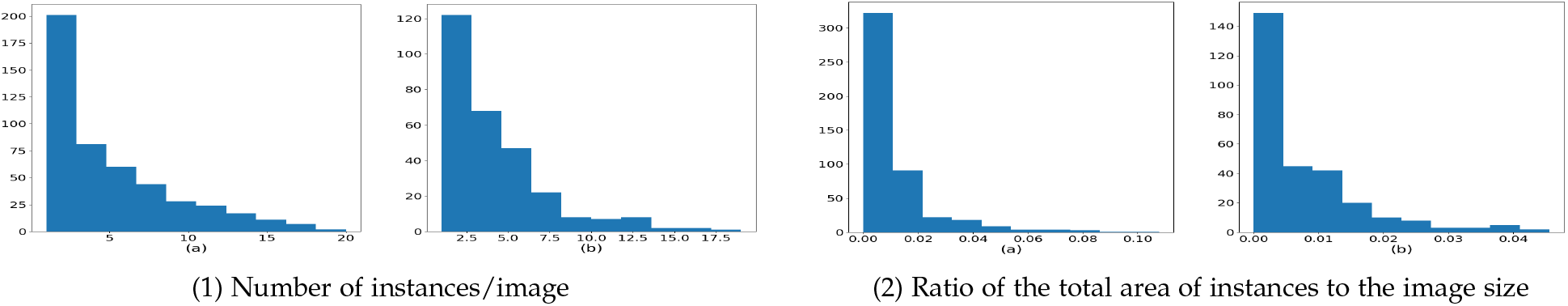
Distribution of the COVID correlates in the segmentation data. Column (a): Ground Glass Opacity, Column (b): Consolidation. In 1.1, we plotted the number of separate GGO and C occurrences in each image. The absolute majority of images have a small number (< 5 occurrences of each type). Histograms in Figure 1.2 complements this finding with the area of occurrences: the absolute majority of them are very small: GGO are < 2% of the image size and C are < 1%. This means that the absolute majority of CT scans has a small number of small occurrences.

### 2.2 Classification

To compare our model to COVIDNet-CT we used the dataset labelled at the image level provided by the same source, [ZLS^+^20], http://ncov-ai.big.ac.cn/download and the split, COVIDx-CT that was used to train COVIDNet-CT model (https://github.com/haydengunraj/COVIDNet-CT), which is publicly available. In total 104900 images from the CNCB dataset were partitioned into 60% training, 20% validation and 20% test data. The difference between COVIDx-CT and the source data is that for COVID and pneumonia classes, only scans with observable infected regions were selected [GWW20].

One of the advantages of our model is a small dataset used for training. We extracted randomly 3000 images from COVIDx-CT training data (1000/class), while maintaining the full size of the validation (21036 images) and test (21182 images) for direct comparison. In the validation split, the shares or Normal/Pneumonia/COVID classes are 43%/35%/22%, in the test split they are 45%/35%/20%.

## 3 Our Approach

Our solution is split into two stages: first, we train an instance segmentation model (Figure 3.1) to predict masks of GGO and C areas. After validation, this model is augmented with a classification module *S* (Figure 3.3) that uses ranked bounding box predictions to classify the whole input image (Figure 3.2).

### 3.1 Segmentation model

Faster R-CNN[RHGS15] and Mask R-CNN [HGDG17] extract regional features from one of the backbone feature maps in three steps: 1) align the coordinates predicted by RPN to the feature map, 2) crop them, 3) resize (RoI align) to the predefined size. As a result, all RoIs are of the same size: *C* × *H* × *W* (*C*: number of maps, *H, W*: height and width of the map). A sample of positive and negative RoIs is constructed to predict the class and refined coordinates of the object. The object’s mask is predicted independently of other objects and classes, by comparing the mask’s logits to the ground truth mask. We train Mask R-CNN to construct the model that is capable of finding a number of small objects of varying shapes, which are widespread in CT scans of patients with COVID, see Figure 1. Most anchor sizes are small (< 32 × 32 pixels) and have a large number of scales (6 in total between 0.1 and 2), allowing for accurate detection of various shapes of GGO and C. For the explantion of the details of the model’s hyperparameters (Non-max suppression, RPN/RoI batch size, foreground and background selection thresholds, etc) see [HGDG17] and our implementation. Examples of segmentation model’s outputs are presented in Figure 2. We use Torchvision implementation of Mask R-CNN https://github.com/pytorch/vision/tree/master/torchvision/models/detection with 5 loss functions: binary cross-entropy for class and Smooth1Loss for bounding box coordinates in RPN, multilabel cross-entropy for class and Smooth1Loss for bounding box coordinates in RoI and pixel-wise class-conditional binary cross-entropy for masks.

**Figure 2:**
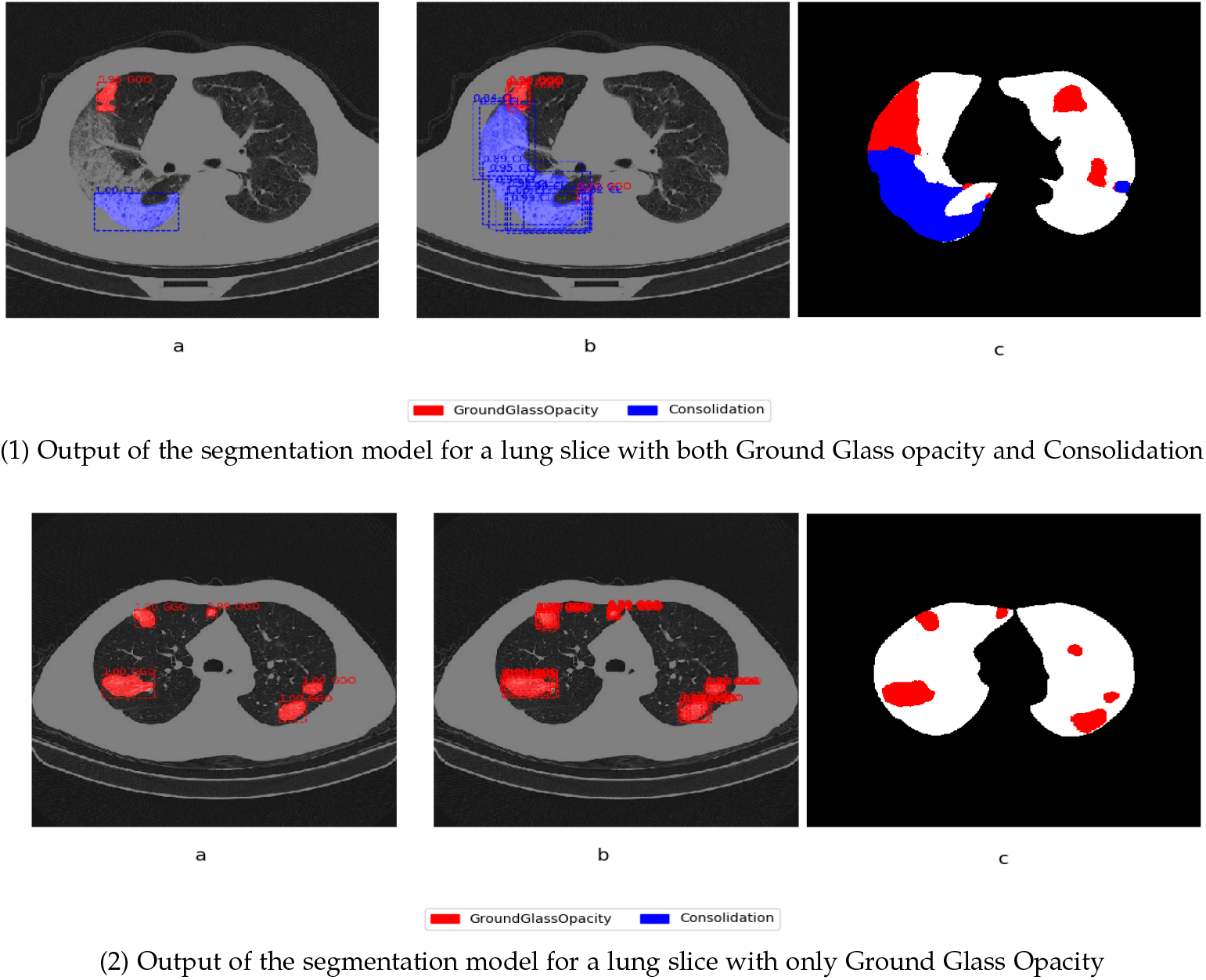
Ground Glass Opacity and Consolidation segmentation predicted by the same model with RoI Non-Max Suppression Threshold=0.25 in column (a) and RoI Non-Max Suppression Threshold=0.75 in column (b). Column (c) is the ground-truth mask. Predictions with scores above RoI score_*θ*_ = 0.75 for each detection and all pixel-level mask logits > 0 are considered positive.

#### Algorithm 1: COVID-CT-Mask-Net algorithm.

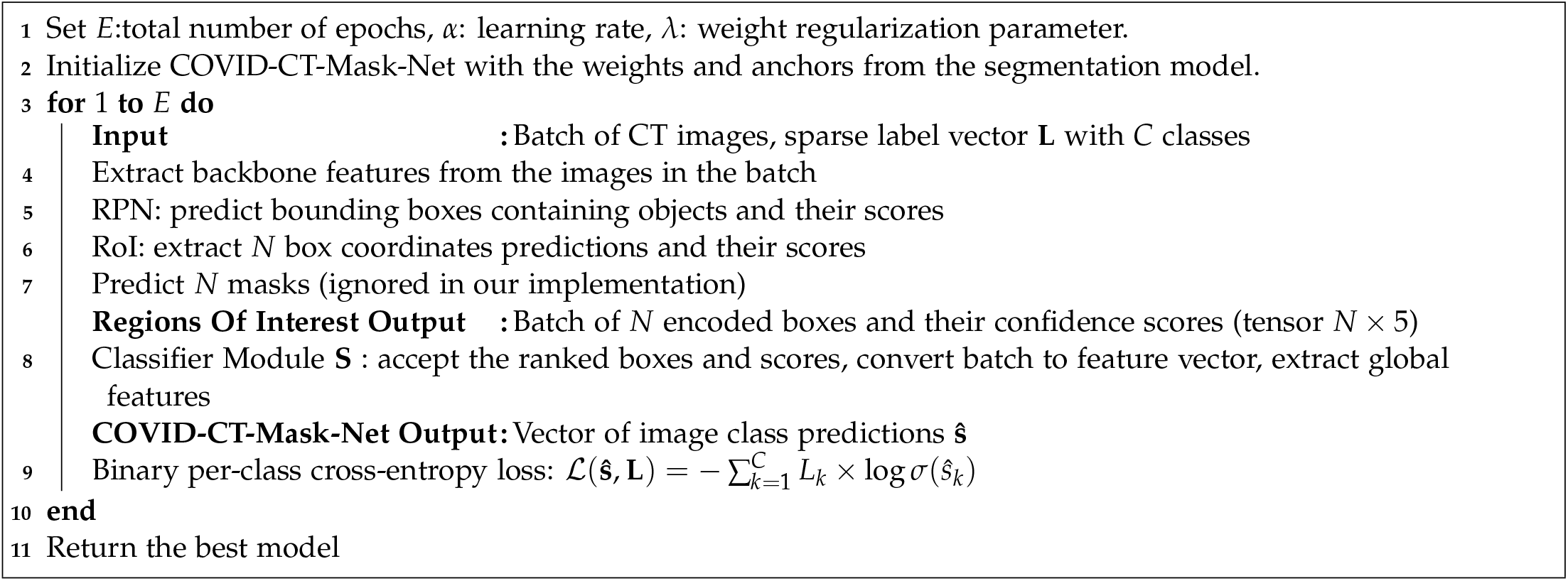

### 3.2 COVID-CT-Mask-Net

We augment Mask R-CNN with a classification module *S* that makes predictions about the whole image. Details of the COVID-CT-Mask-Net algorithm are presented in Algorithm 1 and Figure 3.2. The details of module *S* are in Figure 3.3.

**Figure 3:**
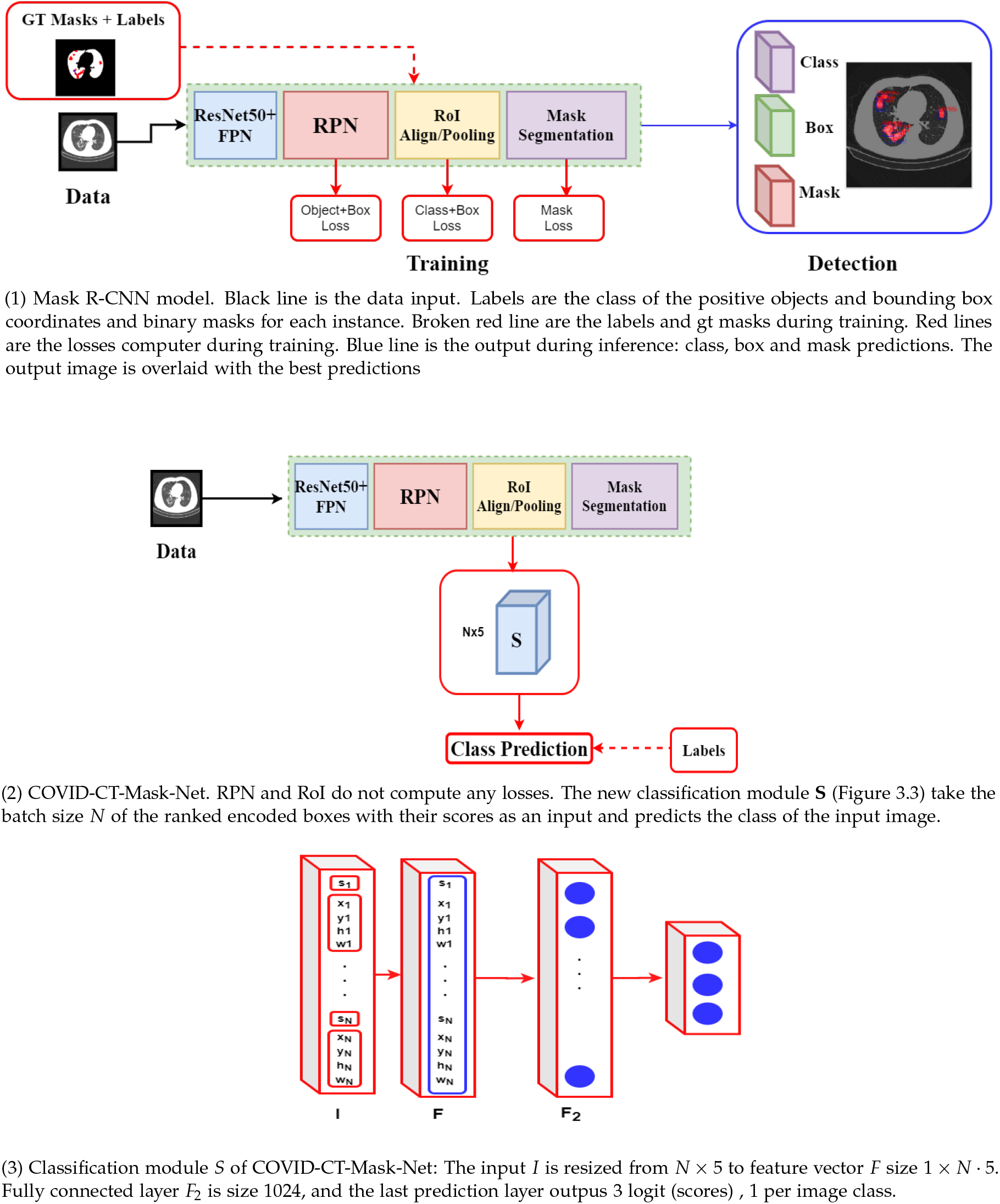
Architecture of the segmentation model, COVID-CT-Mask-Net and classification module *S*.

#### Batch to features

One of the most important steps in Mask R-CNN is construction of the batch in the image by taking a sample of positive (score > *θ*_pos_) and negative (score < *θ*_neg_) RoIs at the training stage. At the inference stage, each RoI predicts a number of encoded bounding boxes (one per class), each with a confidence score, from which, after discarding predictions with scores below score_*θ*_, and overlapping predictions, a batch of *atmostN* highest-scoring bounding box predictions is extracted. Mask R-CNN encodes coordinates to make predictions independent of the images size, so this is a form of normalization.

We transform this process for the purpose of whole image classification, as we need low-scoring regions too, to give the classifier sufficient information, especially for negative images without any GGO and C conditions. To obtain a fixed-size output from RoI stage, we set the score_*θ*_ = − 0.01, so that even very low-scoring predictions are accepted, and RoI output size is fixed to *N* × 5 (*N* encoded bounding box coordinates+confidence score). Normally, RoI decodes these bounding box coordinates by scaling them to the size of the input image, and ranks them based on the confidence score. We ignore the scaling to the image size and use these ranked encoded coordinates with their score as an input in the classification module *S*. The advantage of this approach is that, even if the highest score is very low (in negative images), the predicted coordinates are still ranked (highest to lowest). This ranking pattern is something the classifier *S* can learn. *S* resizes this input into a single feature vector size 1 × (*N* · 5), which maintains the rank of detections. After some filtering and feature extraction, the module predicts the scores for the whole image (COVID, pneumonia, control), see Figure 3.3.

#### Non-maximum suppressions (NMS)

NMS is the threshold value for discarding predictions of the object of the same class. Setting it high means allowing a larger number of predictions in the training sample with IoU> pre-defined NMS threshold. We established that the model learns that overlapping (adjacent) regions with high scores are associated with higher probability of presence of COVID, and hence it improves sensitivity at the cost of lower overall accuracy. To over come this fact, since in many scans GGO or C areas can be very small, and hence produce only one or very few high-scoring box predictions, we set the NMS threshold to 0.75 in both models, thus increasing the sensitivity to COVID.

## 4 Experiments

For COVID-CT-Mask-Net, we re-implement Torchvision’s Mask R-CNN library. During the training of the classifier, RPN and RoI do not compute any loss. The object threshold RoI score_*θ*_ is set to −0.01 to accept all box predictions, even with low scores, to guarantee the batch size and the feature vector remain the same in *S*. We train COVID-CT-Mask-Net in three different ways (Table 1): only classification module *S*, module *S*+batch normalization layers, and full model. To train the full model, a large hack had to be applied both to the RPN and RoI modules: all layers in these modules were set to the training mode, the weights made trainable, and loss computation and all related sampling operations were switched off. Therefore, although formally Mask R-CNN layers were in the evaluation mode, in fact they were updated. Compared to other models, we use a small fraction of the dataset of COVIDx-CT for training, while maintaining the full size of the test and validation sets. As a result, the test/train splits ratio is 7.06, which is the new state-of-the-art, and demonstrates the ability of COVID-CT-Mask-Net to generalize to the unseen data. We use Adam optimizer, learning rate 1*e* − 5, weight regularization parameter 1*e* − 3, and train each algorithm for 50 epochs. For other details of the segmentation algorithm and COVID-CT-Mask-Net see our implementation, https://github.com/AlexTS1980/COVID-CT-Mask-Net.

**Table 1:** Comparison of models’ sizes and data splits used to training, validation and testing.

| Model | Total #parameters | #Trainable parameters | Training | Validation | Test | Ratio Test/Train |
| --- | --- | --- | --- | --- | --- | --- |
| Mask R-CNN (segmentation) | 31.78M | 31.78M | 600 | 150 | - | - |
| COVID-CT-Mask-Net (heads) | 34.14M | 2.25M | 3K | 20.6K | 21.1K | 7.06 |
| COVID-CT-Mask-Net (heads+BN) |  | 2.36M |  |  |  |  |
| COVID-CT-Mask-Net (full) |  | 34.14M |  |  |  |  |
| COVIDNet-CT [GWW20] | 1.8M | 1.8M | 60K | 20.6K | 21.1K | 0.353 |
| COVNet [LQX<sup>+</sup>20] | 25.61M | 25.61M | 3K | 370 | 438 | 0.129 |
| ResNet18 [BGCB20] | 11.69M | 11.69M | 528 |  | 90 | 0.17 |

To evaluate each model, we compute the sensitivity/recall and precision/positive predictive value (PPV) for each class *C* and the overall accuracy of the model:

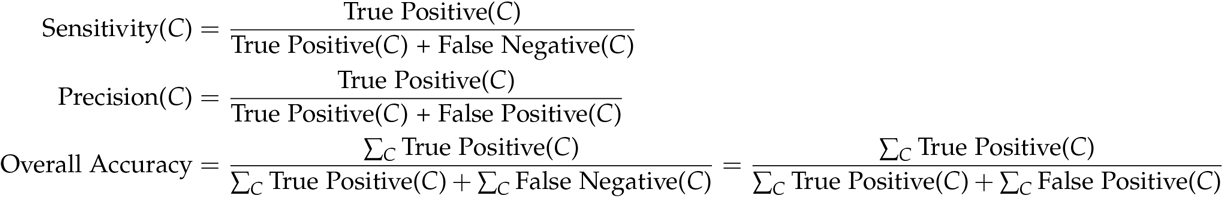

Best results for each trained version of COVID-CT-Mask-Net are presented in Table 2. The model with the classifier head + batch normalization layers produces precision > 90% across all classes. Comparison of our results to other COVID CT detectors for 3 classes is presented in Tables 1 and 3. For COVIDNet-CT we used the best reported model (COVIDNet-CT-A), COVNet and [BGCB20] report only one model.

**Table 2:**
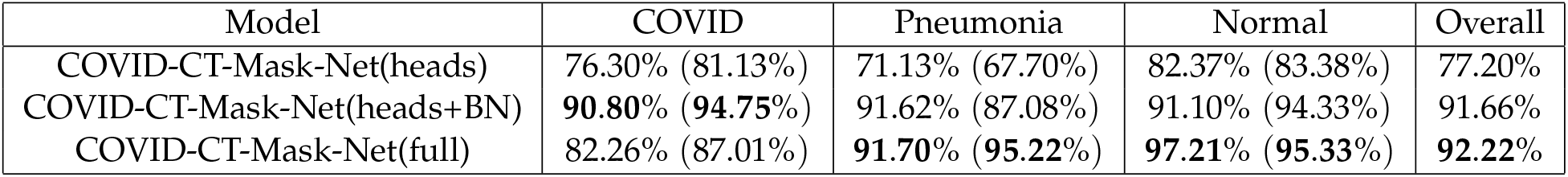
Sensitivity (precision) and overall accuracy results on COVIDx-CT test data (21182 images)

| Model | COVID | Pneumonia | Normal | Overall |
| --- | --- | --- | --- | --- |
| COVID-CT-Mask-Net(heads) | 76.30% (81.13%) | 71.13% (67.70%) | 82.37% (83.38%) | 77.20% |
| COVID-CT-Mask-Net(heads+BN) | **90.80%** (**94.75%**) | 91.62% (87.08%) | 91.10% (94.33%) | 91.66% |
| COVID-CT-Mask-Net(full) | 82.26% (87.01%) | **91.70%** (**95.22%**) | **97.21%** (**95.33%**) | **92.22%** |

**Table 3:** Comparison to other models. The results for COVIDNet-CT were obtained by running the publicly available model (https://github.com/haydengunraj/COVIDNet-CT) on the same test split, results for the other two models are taken from the publication. Last column is the share of COVID observations in the test split. Test split for COVNet has 438 images, ResNet18 90 images.

| Model | COVID Sensitivity | Overall accuracy | COVID prevalence |
| --- | --- | --- | --- |
| Ours | 90.80% | 91.66% | 20% |
| COVIDNet-CT [GWW20] | 92.49% | 97.57% | 20% |
| COVNet [LQX<sup>+</sup>20] | 90.00% | 89.04% | 30% |
| ResNet18 [BGCB20] | 81.30% | 86.70% | 35.79% |

## 5 Conclusions

It is often a challenge to find a sufficiently large dataset to train models for accurate predictions of COVID. One of the strongest features of COVID-CT-Mask-Net’s methodology is the ability to train on very small amounts of data without any balancing and augmentation tweaks. We trained our model on less than 5% of COVIDx-CT training split, and evaluated it on more than 21000 test images achieving a 91.66% overall accuracy and 90.80% COVID sensitivity. The model can be easily and quickly finetuned to new CT data to achieve high COVID detection rate. The source code with all models and weights are on https://github.com/AlexTS1980/COVID-CT-Mask-Net.

## Data Availability

https://github.com/AlexTS1980/COVID-CT-Mask-Net

